# Genome-wide Association Study Identifies *SORCS3* as a Novel Susceptibility Locus for Panic Disorder in the FinnGen Study

**DOI:** 10.1101/2025.07.03.25330813

**Authors:** Joonas Naamanka, Elisa Tasanko, Kalevi Trontti, Bozidar Novak, Veijo Kinnunen, Jaana Suvisaari, Tiina Paunio, Mariliis Vaht, Wei Zhou, Mitja Kurki, Jessica Reinhart, Juliane Boschet-Lange, Roxana Pittig, Andre Pittig, Zuzanna Misiewicz, Manuel Mattheisen, Sandra Meier, Elisabeth B. Binder, Andres Metspalu, Mark Daly, FinnGen, Christoph W. Turck, Angelika Erhardt-Lehmann, Jaana A. Hartiala, Iiris Hovatta

## Abstract

Panic disorder is an anxiety disorder with poorly understood etiology. Although twin studies suggest modest heritability (∼40%), few genetic variants have been associated with it. We carried out a genome-wide association study in the Finnish longitudinal health register-based FinnGen study to identify genetic variants that predispose to panic disorder. FinnGen cases (N=3,549) were defined as individuals with an ICD-10 or ICD-9 lifetime diagnosis of panic disorder. Control subjects (N=159,869) were free of any psychiatric diagnoses. We identified a locus on chromosome 10q25.1 within *SORCS3* that was significantly associated with panic disorder. The minor allele (T; frequency 21%) of the lead variant rs902306 increased the risk of panic disorder by 22% (OR=1.22, 95% CI=1.15-1.30, p-value=1.1×10^−10^). We also investigated serum SORCS3 protein levels in 107 panic disorder cases with or without agoraphobia and 95 controls free of axis I psychiatric disorders collected at the Anxiety Disorders Outpatient Unit of the Max Planck Institute of Psychiatry. Serum SORCS3 levels were 41% higher in panic disorder cases compared to controls (ß=0.694, SE=0.141, p-value=8.7×10^−07^). This finding replicated in 84 subjects from an independent German clinical panic disorder sample (ß=1.137, SE=0.532, p-value=0.04), but not in plasma samples of Finnish panic disorder patients from a biobank. Overall, *SORCS3* is a novel panic disorder locus, which has previously been associated with other psychiatric and neurodevelopmental disorders. *SORCS3* belongs to the sortilin family, with multiple functions related to brain plasticity. Characterization of its role in panic disorder will increase understanding of the neurobiological mechanisms involved in anxiety.

## Introduction

Panic disorder is defined by recurrent, unexpected panic attacks, wherein at least one panic attack must be followed by at least one month of persistent concern about having more attacks, worry about the consequences of the attacks, or maladaptive behaviour (e.g., avoidance of work or school activities) related to the attacks ^1^. The lifetime prevalence of panic disorder is 1.7-3.7% ^2–4^ and it is twice as common in females compared to males ^5^. The age of onset is often in early adulthood, and about 80% of cases have a comorbid mental disorder, most often another anxiety or a mood disorder ^2^. Like in most psychiatric disorders, the etiology is complex, consisting of multiple genetic and environmental risk factors.

A meta-analysis of twin studies has estimated panic disorder heritability to be ∼43% ^6^. Panic disorder has a shared genetic component with other anxiety disorders ^7–9^, and different anxiety disorders often aggregate within individuals and families ^6^. Therefore, most previous genome-wide association studies (GWAS) have investigated panic disorder as part of a larger composite phenotype of anxiety disorders ^7, 8, 10, 11^. Of the panic disorder specific GWAS, an early study in 200 Japanese cases and 200 controls identified two significant loci, one within the *PKP1* (p-value=4.6×10^−08^) and another within *TMEM16B* (p-value=3.7×10^−09^) ^12^. SNPs within *TMEM132D* were associated with panic disorder (p-value=5.1×10^−07^) in a German study of 216 cases and 222 controls ^13^, and this finding was replicated in a multi-ethnic sample of 1,670 cases and 2,266 controls and validated in a mouse model ^13, 14^. The largest GWAS meta-analysis of panic disorder to date, with 2,147 cases and 7,760 controls of European origin, did not detect genome-wide significant associations ^15^.

To examine the genetic architecture of panic disorder, we took advantage of the large Finnish register-based sample from the FinnGen (http://finngen.fi), which combines healthcare register data with genetic information, and conducted a GWAS. We then sought to replicate our finding in other populations, using summary level data from the UK Biobank and Estonian Biobank. Finally, to explore the potential as a measurable biomarker for panic, we examined serum protein levels of SORCS3, our most significant finding from the FinnGen GWAS, in two German clinical panic disorder samples and a Finnish biobank sample.

## Materials and Methods

### FinnGen Study Sample

The FinnGen (https://www.finngen.fi/en) study combines genotype data with longitudinal health register data of Finland. Panic disorder cases were defined according to ICD-10 F41.0 or ICD-9 3000B, classified to mild, moderate, or severe panic disorder (F41.08, F41.00, F41.01), or unspecified panic disorder (F41.09) (**Table 1**). We excluded cases with psychotic disorders, autism, or intellectual disability. Exclusion criteria for controls included history of any psychiatric endpoint. Additionally, we adjusted the age range of controls to match that of cases. Based on this approach, we included 3,549 cases and 159,869 control subjects in this study (see **Supplemental Methods** for details).

**Table 1.** Clinical characteristics of the FinnGen Study.

|  | All | Females | Males |
| --- | --- | --- | --- |
| Panic disorder |  |  |  |
| Cases | 3 549 | 2 592 | 957 |
| Controls | 159 869 | 86 937 | 72 932 |
| Mean year of birth |  |  |  |
| Cases | 1970 (16.8) | 1972 (16.8) | 1967 (16.3) |
| Controls | 1959 (17.1) | 1961 (17.1) | 1956 (16.7) |
| **<sup>a</sup>Panic disorder diagnoses** |  |  |  |
| Panic disorder (F41.0) | 3 467 | 2 550 | 917 |
| <sup>b</sup>Mild panic disorder (F41.08) | 151 | 118 | 33 |
| <sup>b</sup>Moderate panic disorder (F41.00) | 1 046 | 771 | 275 |
| <sup>b</sup>Severe panic disorder (F41.01) | 272 | 198 | 74 |
| Unspecified panic disorder (F41.09) | 1 854 | 1 371 | 483 |
| Panic disorder (3000B) | 85 | 46 | 39 |
| Broad definition of panic disorder (3002B) | 31 | 11 | 20 |
| **Psychiatric comorbidities among cases** |  |  |  |
| No psychiatric comorbidities | 628 | 434 | 194 |
| <sup>c</sup>Major depressive disorder (MDD) | 2 023 | 1 530 | 493 |
| <sup>d</sup>Generalized anxiety disorder (GAD) | 524 | 402 | 122 |
| <sup>e</sup>Other anxiety or stress disorder | 2 208 | 1 671 | 537 |

Data is shown as number of subjects or year of birth (SD).
<sup>a</sup>Panic disorder cases were defined as follows: ICD-10 F41.0, F41.08, F41.00, F41.01, F41.09; ICD-9 3000B, 3002B. Control subjects were free from any psychiatric diagnosis.
<sup>b</sup>Severity of panic disorder was defined as mild: <4 panic attacks within a month; moderate: ≥4 panic attacks within a month; severe: ≥4 panic attacks within a week.
<sup>c</sup>MDD cases were defined as follows: ICD-10 F32, F33; ICD-9 2961; ICD-8 2980, 3004, 79020.
<sup>d</sup>GAD cases were defined as follows: ICD-10 F41.1; ICD-9 3000C.
<sup>e</sup>Other anxiety or stress disorder cases were defined as follows: ICD-10 F40.00, F40.01, F40.1, F40.2, F40.8, F40.09, F41.1, F41.2, F41.3, F41.8, F41.9, F42, F43, F44, F45, F48; ICD-9 3000A, 3000C, 3001, 3003, 3004, 3006, 3007, 3008; ICD-8 30000, 3001, 3003, 3004, 3005, 3006, 3007, 3008, 3009.

### Max Planck Institute of Psychiatry Study Sample

Panic disorder patients were recruited in the Anxiety Disorders Outpatient Unit at the Max Planck Institute of Psychiatry (MPIP). DSM-IV panic disorder with and without agoraphobia was the primary diagnosis. Panic disorder due to a medical or neurological condition or the presence of a comorbid Axis II disorder was an exclusion criterion. Control subjects were recruited from a Munich-based community sample and screened for the absence of axis I psychiatric disorders. The analysed sample consisted of 107 patients (52 males and 55 females) and 99 controls (51 males and 48 females)

An independent depression sample (58 male and 62 female patients and 40 male and 40 female controls) was recruited at the MPIP within the Munich Antidepressant Response Study (MARS) ^16, 17^. Diagnosis was ascertained according to the DSM-IV criteria. The exclusion criteria were alcohol or substance abuse or dependence, comorbid somatization disorder, and depressive disorders owing to general medical or neurologic conditions. Controls were recruited and characterized as described for panic disorder (see **Supplemental Methods** for details).

### Würzburg Study Sample (WU)

Patients with anxiety disorders aged between 18 and 65 years were recruited via the outpatient clinic of the University of Würzburg, local advertisement, and referral. Inclusion criteria required a primary diagnosis of panic disorder. Exclusion criteria comprised acute suicidality, psychosis, bipolar disorder, borderline personality disorder, substance dependence, intellectual disability, any severe acute medical condition, pregnancy, medical advice to avoid stressful situations, and any contraindication for exposure therapy ^18^. Diagnostic interview for mental disorders was conducted using the Mini-DIPS according to DSM-5 ^19^. Controls were recruited within the MEGA TRR 58 study (‘Fear, Anxiety, Anxiety Disorders’, project Z02/4) and screened for absence of any mental illness. Serious neurological and somatic illnesses as well as pregnancy were exclusion criteria. Blood samples, including serum, were processed immediately after collection and stored at -80°C. The analyzed sample included 19 cases (14 females and 5 males) and 65 controls (46 females and 19 males) (see **Supplemental Methods** for details).

### Helsinki Biobank Study Sample

Plasma samples from 300 panic disorder patients (58 males, 242 females), 300 patients with major depressive disorder (MDD; 54 males, 246 females), and 300 controls (58 males, 242 females) were obtained from the Helsinki Biobank. Inclusion criterion for panic disorder was ICD-10 F41.0 diagnosis and exclusion criteria psychotic disorders (F20-F29), autism (F84.0, F84.1, F84.5), or intellectual disability (F70-F79). Inclusion criterion for MDD was ICD-10 F33 diagnosis and exclusion criteria psychotic disorders (F20-F29), autism (F84.0, F84.1, F84.5), or intellectual disability (F70-F79) and panic disorder. Additionally, we required that the plasma sample was taken after the first diagnosis (the average time between the first diagnosis of panic disorder or MDD and blood sampling was 6.8 years). Controls were required to have no history of any psychiatric endpoint.

### GWAS for Panic Disorder

Subjects from the FinnGen were genotyped with Illumina and Affymetrix arrays (Illumina Inc., San Diego, and Thermo Fisher Scientific, Santa Clara, CA, USA) as described (https://www.finngen.fi/en/researchers/genotyping). Genotyping and imputation with the Finnish population-specific SISu v3 reference panel were conducted by the FinnGen, as described (https://www.protocols.io/view/genotype-imputation-workflow-v3-0-xbgfijw). Briefly, ∼16 million single nucleotide polymorphisms (SNPs) were available. After including SNPs with minor allele frequency (MAF) ≥0.01 that exceeded the quality control criteria (Info score>0.8), 9,362,465 genetic variants spanning autosomal and X chromosomes were available for GWAS in 163,418 individuals. GWAS was performed using the Scalable and Accurate Implementation of GEneralized mixed model (SAIGE) v0.20 ^20^ with a kinship matrix as a random effect and age, sex, the first 10 principal components (PCs), and genotyping batch as fixed effects. The dataset uses genome build 38 (hg38).

### Prioritizing Variants at the *SORCS3* Locus

To functionally validate and prioritize variants at the *SORCS3* locus, we first used the Functional Mapping and Annotation of Genome-wide Association Studies (FUMA GWAS) pipeline ^21^. Independent SNPs from GWAS summary results were identified as variants with a p-value<5.0×10^−08^ and moderately low linkage disequilibrium (LD) with each other (r2<0.2). As part of FUMA, we also performed gene set analysis using the MAGMA tool ^22^.

### Comparison of Association Signals at the *SORCS3* Locus with Panic Disorder and Related Psychiatric Outcomes

The *SORCS3* locus has previously been associated with other psychiatric conditions, including a composite phenotype of anxiety disorders ^23^. To further validate that the association signals with our lead SNPs were deriving from panic disorder, we performed comparative association analyses with our lead *SORCS3* variants (rs902306 and rs1021362) and other psychiatric comorbidities, such as GAD and phobias; and MDD in the FinnGen sample. Controls for all comparative analyses remained the same as used in the GWAS for panic disorder. First, we tested *SORCS3* SNP associations with strict panic disorder excluding all cases with comorbid psychiatric endpoints, such as other anxiety disorders, MDD, or substance use disorder. To examine whether *SORCS3* association for panic disorder may arise due to an overlapping diagnosis with other comorbidities, we next performed analysis for MDD or GAD/phobia with our lead variants where panic disorder was excluded from the case definition. All analyses were performed using SAIGE v0.20 ^20^ with a kinship matrix as a random effect and age, sex, first 10 PCs, and genotyping batch as fixed effects in the model.

### Replication Analyses in the UK Biobank and Estonian Biobank

To investigate whether the association of our *SORCS3* lead SNPs replicates in other studies, we acquired summary level data for panic disorder with *SORCS3* lead variants (rs902306 and rs1021362) from the UK Biobank and the Estonian Biobank (see **Supplemental Methods** for a description of the samples).

### Genetic correlation with other psychiatric phenotypes

To estimate the SNP-based heritability of panic disorder and its genetic correlations (rG) with other psychiatric or cognitive phenotypes we used the LD Score Regression (LDSC) software ^24, 25^, which uses the linkage disequilibrium (LD) score regression method. The LD scores were computed from European ancestry subset of the 1000 Genomes dataset ^26^ and only the SNPs in HapMap 3 ^27^ were included to ensure high quality imputation.

We examined the genetic correlations of panic disorder with psychiatric or cognitive phenotypes where GWAS summary statistics from the Psychiatric Genomics Consortium (https://pgc.unc.edu/for-researchers/download-results/) or Complex Trait Genetics lab at the Center for Neurogenomics and Cognition Research in Amsterdam (https://ctg.cncr.nl/software/summary_statistics) were available. When multiple summary statistics were available for one phenotype, we chose the one with the largest sample size. Genetic correlation calculations were performed for major depressive disorder ^28^, anxiety factor score ^7^, attention deficit/hyperactivity disorder (ADHD) ^29^, cannabis use disorder (CUD) ^30^, schizophrenia ^31^, insomnia ^32^, bipolar disorder ^33^, Tourette’s syndrome ^34^, obsessive-compulsive disorder (OCD) ^35^, autism spectrum disorder ^36^, anorexia nervosa ^37^, quantitative measures from the Alcohol Use Disorders Identification Test (AUD) ^38^, and intelligence (IQ) ^39^.

### Evaluation of serum or plasma SORCS3 Levels

Serum or plasma SORCS3 levels were measured with the Human VPS10 Domain-Containing Receptor SORCS3 (SORCS3) ELISA Kit (MyBiosource, San Diego, USA) according to manufacturer’s instructions.

### The MPIP sample

The analysis was carried out at the MPIP from serum samples. The panic disorder analysis was carried out in two batches balanced by sex, with 54 patients and 69 controls (batch 1), and 53 patients and 30 controls (batch 2). Four control samples from batch 2 were outside of the detection limit of the kit (0.5-16 ng/ml) and were excluded from the analyses. The MDD analysis was carried out in a third batch, with 120 patients and 80 controls. Obtained SORCS3 concentration values were log-transformed, followed by combined and sex-stratified linear regression analysis with panic disorder or MDD as an exposure variable, after adjustment for age, sex, batch, and project (if applicable). Lastly, an interaction term between panic disorder and sex was added to the model to test whether SORCS3 levels by patient status differ based on sex.

### The WU sample

The analysis was carried out at the University of Helsinki from serum samples. Four samples were below the detection limit of the kit (0.5 ng/ml) and were excluded leaving 84 samples in the final analysis. Betas and standard errors were derived from linear regressions using untransformed serum SORCS3 levels in all subjects. Sex-stratified analyses were carried out with panic disorder as an exposure variable, after adjustment for age, sex (if applicable), and plate. An interaction term between panic disorder and sex was included in the model to test whether SORCS3 levels by patient status differ by sex.

### The Helsinki Biobank sample

SORCS3 levels were determined from plasma samples at the University of Helsinki. According to the manufacturer, the plasma SORCS3 levels are generally lower than that of the serum levels. Consequently, 315 (35%) samples were outside of the detection limit of the kit (0.5-16 ng/ml) and were excluded from further analysis. Linear regression for log-transformed SORCS3 plasma levels was conducted with panic disorder as an exposure variable, after adjustment for age and sex (if applicable) in all subjects (n=401) and separately in females (n=321) and males (n=80). Similarly, linear regression for log-transformed SORCS3 plasma levels was performed where MDD was treated as exposure variable in all subjects (n=378) and sex-stratified analysis including 304 females and 74 males.

### Diagnostic Accuracy

To evaluate the performance of our binary classification model and whether SORCS3 levels can be utilized to diagnose patients with and without the disease, we constructed a receiver operating characteristic (ROC) curve for panic disorder using logistic regression and plotting predicted probabilities with serum SORCS3 levels in the model including age, sex, batch, and project (MPIP); or age, sex and plate (WU) as covariates. In comparison, we also constructed a ROC curve for MDD after fitting SORCS3 levels, age, and sex in the model in the MPIP depression sample.

## Results

### Identification of *SORCS3* as a novel locus for panic disorder

We identified 132 genome-wide significant (p-value<5.0×10^−08^) SNPs through a GWAS for panic disorder in the FinnGen (**Figure 1**). All variants localized in chr10q25.1 in introns 1 and 2 of the *sortilin related VPS10 domain containing receptor 3* (*SORCS3*) gene (**Figure 1**). Minor allele T of our lead variant rs902306 was associated with a 22% increased risk of panic disorder among 3,549 cases and 159,869 controls free from any psychiatric end points (OR=1.22, 95% CI=1.15-1.30, p-value=1.1×10^−10^) (**Table 2**).

**Figure 1.**
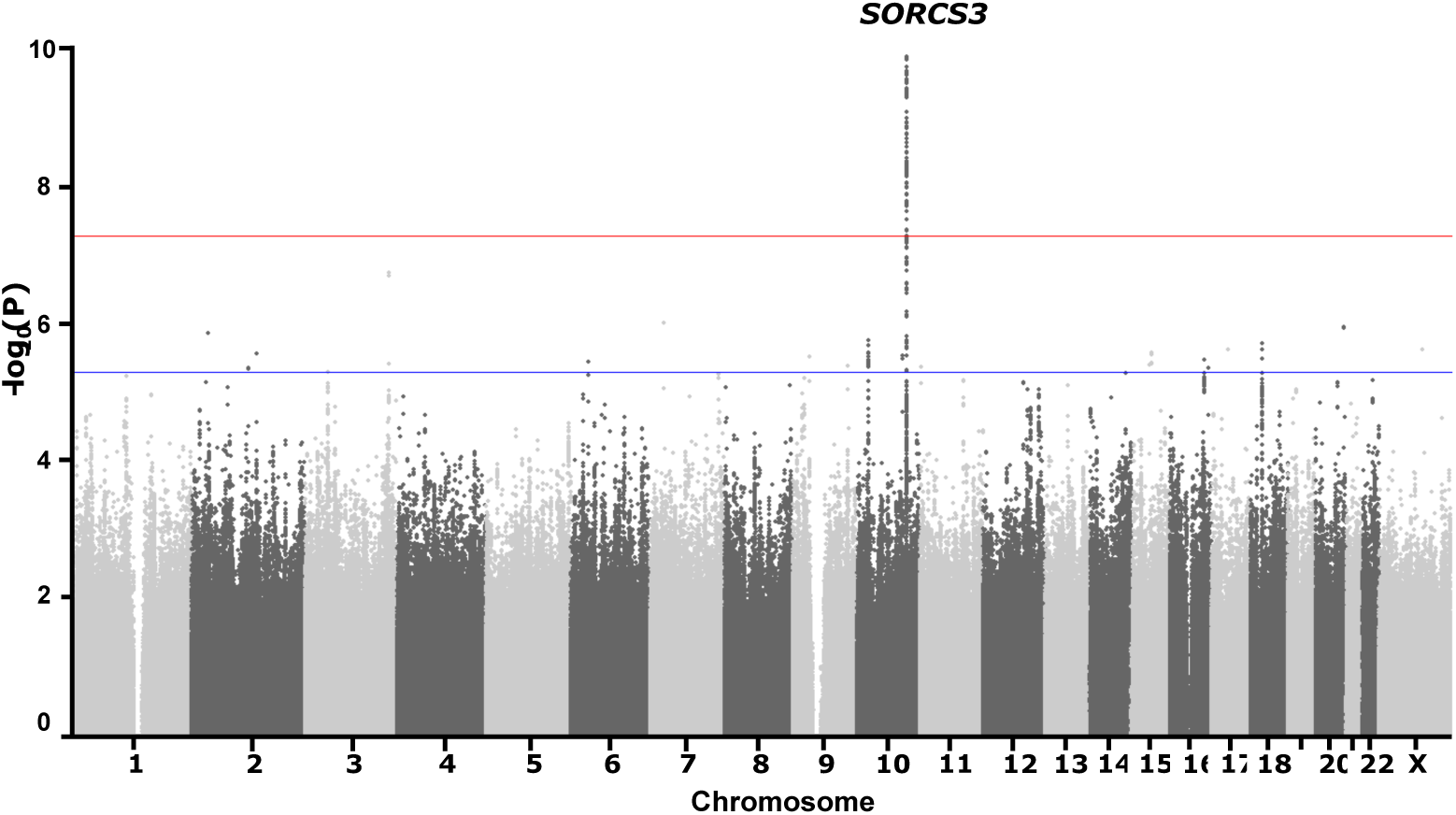
Manhattan plot of GWAS analysis for panic disorder. Genome-wide thresholds for significant (p-value=5.0×10^−08^) and suggestive (p-value=5.0×10^−06^) associations are indicated by the horizontal red and blue lines, respectively. A novel locus within *SORSC3* on chromosome 10q25.1 was significantly associated with panic disorder in subjects from the FinnGen.

**Table 2.** *SORCS3* Lead SNP Associations for Panic Disorder in the FinnGen.

|  |  |  | **rs902306** |  | **rs1021362** |  |
| --- | --- | --- | --- | --- | --- | --- |
| Position (hg38) |  |  | chr10:104,772,457 |  | chr10:104,851,510 |  |
| EA/NEA |  |  | **T/C** |  | **G/A** |  |
| EAF |  |  | 0.21 |  | 0.27 |  |
| **Strata** | **Cases** | **Controls** | **OR (95% CI)** | **p-value** | **OR (95% CI)** | **p-value** |
| All | 3 549 | 159 869 | 1.22 (1.15-1.30) | $1.1 \times 10^{-10}$ | 1.20 (1.13-1.27) | $1.2 \times 10^{-10}$ |

EA, effect allele; NEA, non-effect allele, EAF, effect allele frequency; OR, odds ratio; CI, confidence interval.

Although all our significant GWAS associations for panic disorder were localized within *SORCS3*, we utilized FUMA GWAS pipeline ^21^ to identify additional signals in this locus. This analysis yielded one independent significant SNP, rs1021362, where minor G-allele (MAF 27%) was associated with 20% increased risk of panic disorder (OR=1.20, 95% CI=1.13-1.27, p-value=1.2×10^−10^) (**Table 2**). Rs1021362 is located 79 kb downstream from rs902306 and is in moderately low linkage disequilibrium (LD) with our lead SNP (r2=0.29 in Finns, r2=0.19 in all Europeans). As part of FUMA GWAS ^21^, we performed a gene set analysis using MAGMA ^22^, which did not reveal additional genes associated with panic disorder. Moreover, regional plot of the *SORCS3* locus showed two distinct LD clusters, with rs902306 and rs1021362 as the most significant SNPs of these nearby regions (**Figure 2A-B**).

**Figure 2.**
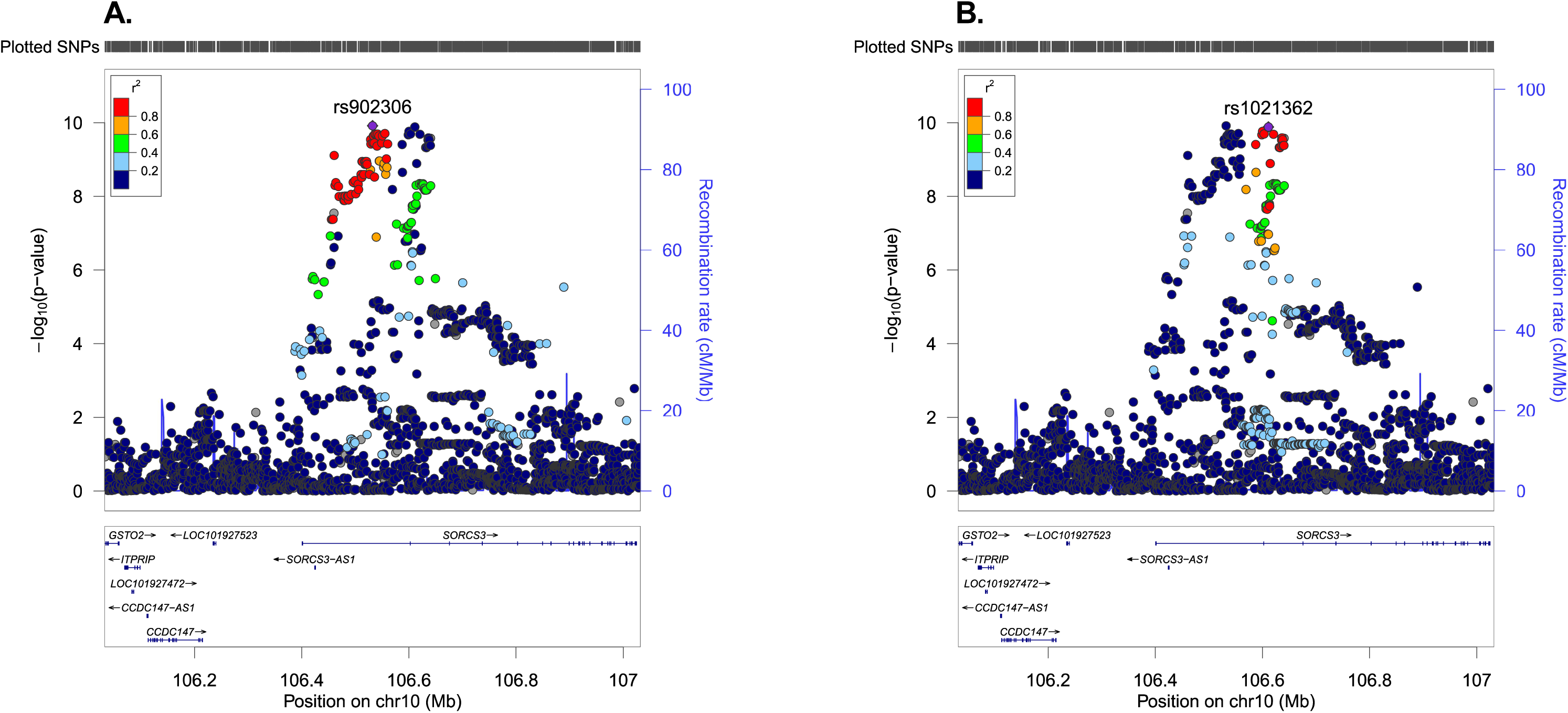
Regional plots of *SORCS3* locus for panic disorder. Each region is centered on the lead SNP (purple diamond) and the genes in the interval are indicated in the bottom panel. The degree of linkage disequilibrium between the lead SNP and other variants is shown as r^2^ values according to the color-coded legend in the box. Regional plots with (**A**) GWAS lead SNP rs902306, and (**B**) an independent significant variant rs1021362 as index SNPs are shown.

Lastly, we performed a phenome-wide association study (PheWAS) with our *SORCS3* variants. We utilized FUMA GWAS ^21^ to identify genome-wide significant (p-value<5.0×10^−08^) associations with our *SORCS3* variants and/or proxy SNPs with high LD (r2≥0.8) with our lead SNPs and other phenotypes. Our lead SNPs have not been significantly associated with any other traits in previous GWAS studies. However, the chr10q25.1 locus has been associated with various traits and psychiatric conditions previously, including a composite anxiety disorder phenotype ^23^ and MDD ^40, 41^ (**Supplementary Table 1**).

### Comparative analyses with other psychiatric disorders

Panic disorder is often comorbid with other psychiatric disorders (**Supplementary Figure 1**). To examine whether our *SORCS3* association can be attributed to panic disorder, we performed comparative analyses in the FinnGen with different panic disorder comorbidities. First, we performed a SNP association with lead *SORCS3* variants for panic disorder after excluding all other psychiatric comorbidities from case-definition (panic disorder only). In this analysis, *SORCS3* variants increased the disease risk by ∼22% (OR=1.20, 95% CI=1.04-1.37, p-value=0.01; OR=1.24, 95% CI=1.10-1.41, p-value=6.3×10^−04^ with rs902306 and rs1021362, respectively) (**Supplementary Table 2**). Effect estimates for panic disorder remained virtually unchanged compared to those without excluded psychiatric comorbidities, but significance was mitigated most likely due to substantial decrease in the number of cases (n=628) (**Supplementary Table 2, Table 2**).

Next, we tested whether *SORCS3* variants were associated with generalized anxiety disorder (GAD) or phobias after exclusion of panic disorder from the case definition. As shown in **Supplementary Table 2**, there was a modest association with rs902306 and risk of GAD/phobia (OR=1.07, 95% CI=1.01-1.13, p-value=0.03), but not with rs1021362. Sample size for GAD/phobia (3,738 cases and 161,438 controls) was similar to panic disorder used in our discovery GWAS (**Supplementary Table 2, Table 2**).

Lastly, we tested whether *SORCS3* variants were associated with MDD. After exclusion of panic disorder diagnosis from the case definition, rs902306 T-allele was associated with 5% increased risk for MDD in 25,906 cases and 161,371 controls (OR=1.05, 95% CI=1.03-1.08, p-value=6.4×10^−05^) (**Supplementary Table 2**). Similarly, rs1021362 G-allele was increasing the risk of MDD by 5% (OR=1.05, 95% CI=1.02-1.07, p-value=6.6×10^−05^) (**Supplementary Table 2**). Although our *SORCS3* variants showed modest association with MDD, the effect estimates and significance were weaker for MDD compared to panic disorder, despite a much larger sample size (**Supplementary Table 2, Table 2**).

### Replication analyses in the UK Biobank and the Estonian Biobank

To investigate whether our *SORCS3* lead SNPs rs902306 and rs1021362 associate with panic disorder in other cohorts, we acquired summary level data from the UK Biobank and the Estonian Biobank. Neither of the SNPs associated with panic disorder in these samples. For example, results from logistic mixed model after adjustment for PC1-10, age, age^2^, sex, age∗sex, and age^2^ sex indicated that our lead SNP rs902306 did not affect the risk of panic disorder in the UK Biobank (OR=1.00, 95% CI=0.92-1.09, p-value=0.99). Similar results were detected in the Estonian Biobank after adjustment for PC1-10, age, and sex (OR=1.02, 95% CI 0.98-1.07, p-value=0.25). (**Supplementary Table 3**).

### Heritability and genetic correlation of FinnGen panic disorder with other psychiatric phenotypes

LD score regression produced a SNP-based heritability estimate of 0.14 on the liability scale, assuming a population prevalence of 3%. The highest genetic correlations were observed with MDD (r_g_=0.76, p-value=4.1×10^−21^) and anxiety factor score (r_g_=0.64, p-value=5.0×10^−04^). The only negative statistically significant correlation estimate was with intelligence (*r*_g_=-0.3, p-value=7.5×10^−09^) (**Figure 3).** All correlations were study-wide significant (Bonferroni corrected threshold 0.05/13 endpoints=3.8×10^−03^) except for Tourette’s syndrome, OCD, anorexia, and AUD.

**Figure 3.**
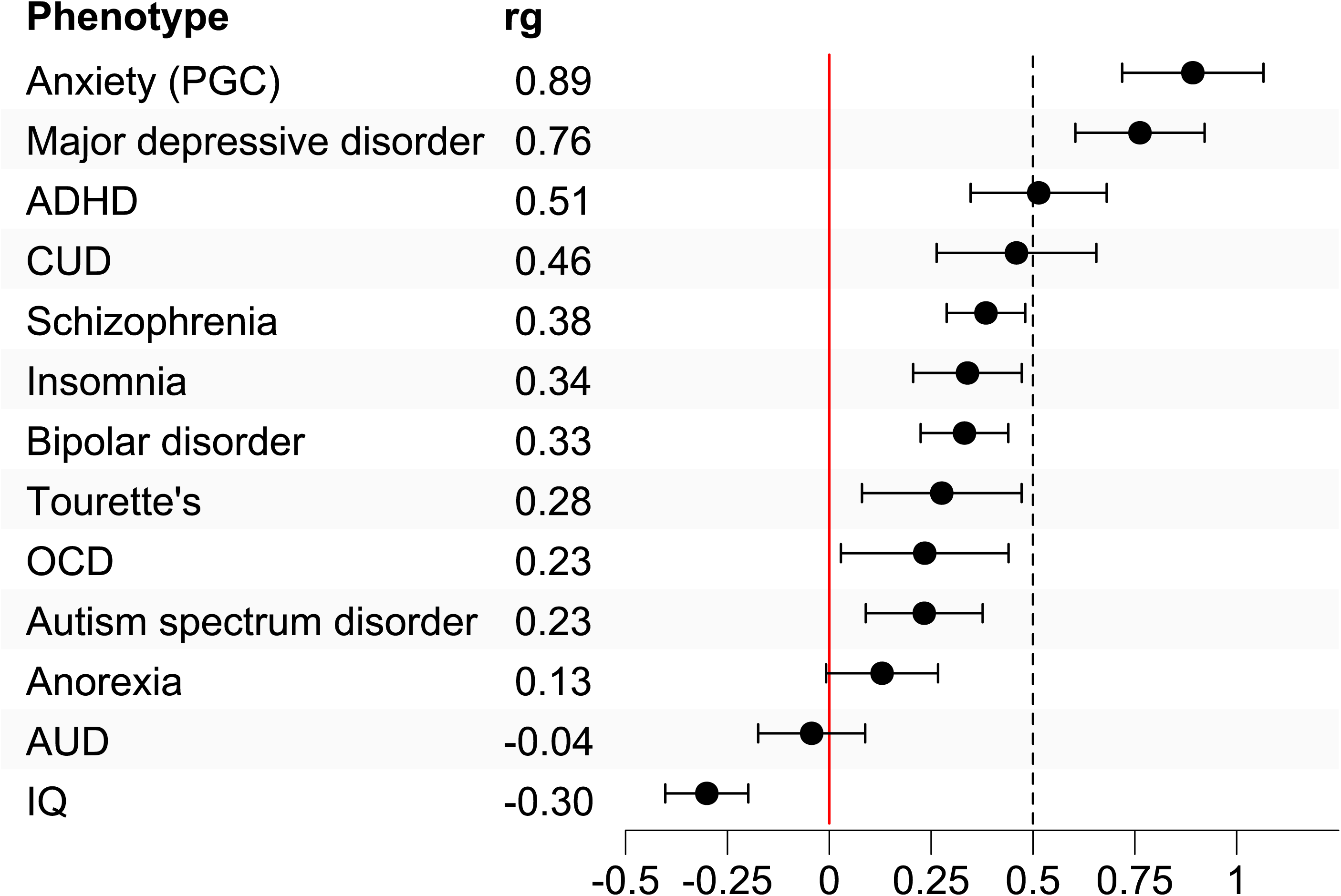
Genetic correlations between panic disorder and other psychiatric or cognitive phenotypes. Genetic correlations between panic disorder and 12 psychiatric phenotypes and intelligence are displayed. The dots represent the point estimates for the correlations and the lines show the 95% confidence intervals. PGC = Psychiatric Genomics Consortium, ADHD = attention deficit/hyperactivity disorder, CUD = cannabis use disorder, OCD = obsessive-compulsive disorder, AUD = quantitative measures from the Alcohol Use Disorders Identification Test, and IQ = intelligence quotient.

### Higher serum SORCS3 levels in panic disorder cases compared to controls

The extracellular domain of the SORCS3 protein can be shed through a regulated proteolytic cleavage and detected in the blood. To investigate whether SORCS3 protein concentrations differ between panic disorder cases and controls, we carried out ELISA using serum from an independent sample of 107 German panic disorder patients and 95 controls, collected at the MPIP. The mean SORCS3 levels were 10.14 ng/ml in male patients, 5.42 ng/ml in male controls, 8.53 ng/ml in female patients, and 6.06 ng/ml in female controls. Overall, panic disorder patients had 41% higher SORCS3 serum levels compared to controls (beta=0.694, SE=0.141, p-value=8.7×10^−07^). In males, the difference was 48.5% (beta=0.874, SE=0.212, p-value=6.4×10^−05^) and in females 31.9% (beta=0.527, SE=0.189, p-value=6.4×10^−03^), although there was no evidence that SORCS3 levels by panic disorder were significantly different between males and females (p-interaction=0.22) (**Figure 4A**). Linear regressions were performed using log-transformation of SORCS3 levels and adjustment for age, sex (where applicable), batch, and project. Furthermore, these results remained virtually unchanged when anxiolytic or antidepressant medication use was added into the model (beta=0.698, SE=0.156, p-value=8.2×10^−06^ in all subjects). In contrast, log-transformed SORCS3 serum levels did not differ between patients with MDD (n=120) and controls (n=80) after adjustment for age and sex among all subjects (beta=0.180, SE=0.100, p-value=0.07) or among males (beta=0.180, SE=0.133, p-value=0.18) and females (beta=0.181, SE=0.149, p-value=0.23) (**Figure 4B**).

**Figure 4.**
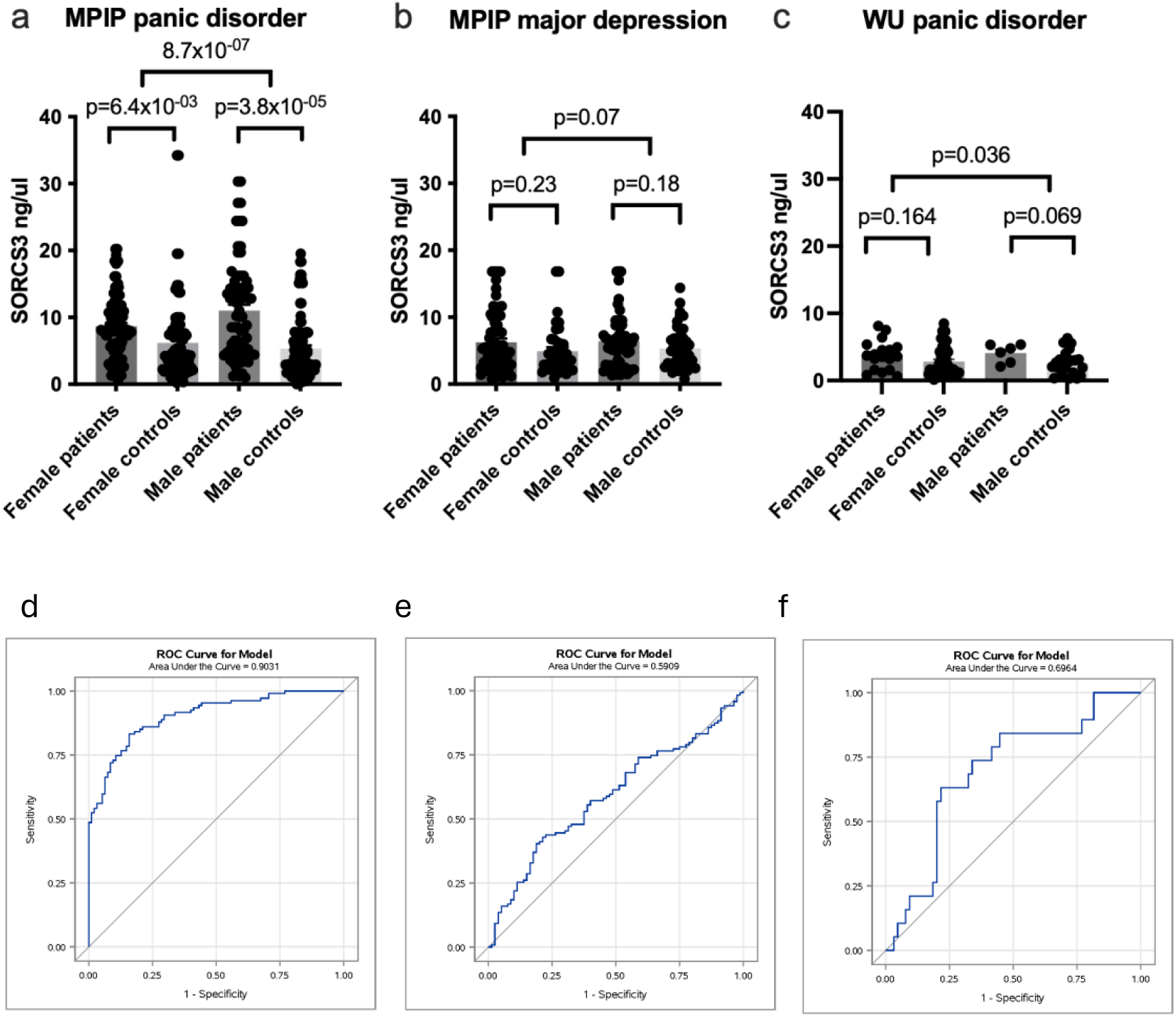
Serum SORCS3 levels in panic disorder and major depressive disorder. We measured SORCS3 protein concentration by ELISA **A.** from 107 panic disorder patients (52 males and 55 females) and 95 controls (49 males and 46 females), **B.** from 120 MDD patients (58 males and 62 females) and 80 controls (40 males and 40 females), and **C.** from 19 panic disorder patients (5 males and 14 females) and 65 controls (19 males and 46 females). Unadjusted SORCS3 levels are shown for males and females separately. P-values are obtained from a linear regression analysis of log-transformed SORCS3 levels after adjustment for age, sex (if applicable), analysis batch, and the project id. Area under the curve from ROC analysis was adjusted for age, sex, analysis batch and project for panic disorder in MPIP (**D**); age and sex for MDD in MPIP (**E**); and age, sex and plate for panic disorder in WU (**F**) sample.

To investigate SORCS3 serum levels in an independent sample, we analyzed samples from 19 cases and 65 controls collected at the WU. The mean SORCS3 levels were 4.47 ng/ml in male patients (n=5), 2.96 ng/ml in male controls (n=19), 4.00 ng/ml in female patients (n=14), and 2.88 ng/ml in female controls (n=46). Overall, panic disorder patients had 30% higher SORCS3 serum levels compared to controls (beta=1.137, SE=0.532, p-value=0.04). In addition, we did not detect significant interaction for panic disorder on SORCS3 levels by sex (p-interaction=0.494) (**Figure 4C**).

Another independent sample utilized was from the Helsinki Biobank, where SORCS3 plasma levels were available in 207 panic disorder cases, 184 MDD cases, and 194 controls. Panic disorder (beta=-0.080, SE=0.068, p-value=0.24) or MDD (beta=-0.069, SE=0.072, p-value=0.34) were not significantly associated with log-transformed SORCS3 levels, and no significant interaction by sex was detected with panic disorder (p-interaction=0.587) or MDD (p-interaction=0.70) (**Supplementary Figure 2**).

To evaluate the performance of our binary classification model for diagnosing patients with and without panic disorder, we performed ROC curve analysis in the MPIP panic disorder sample utilizing logistic regression and plotting predicted probabilities after adjustment for covariates. Among all subjects, 1 ng/ml of serum SORCS3 levels increased the risk of panic disorder by 15% (OR=1.15, 95% CI=1.07-1.24, p-value=1.7×10^−04^) after adjustment for age, sex, batch, and project. The area under the curve (AUC) was 0.90, indicating excellent ability to diagnose patients with and without the disease (**Figure 4D**).

To replicate these findings with panic disorder, we performed logistic regression in the WU sample and plotted predicted probabilities after adjustment for age, sex and plate. Among 84 subjects, 1 ng/ml of serum SORCS3 levels increased the risk of panic disorder by 32% (OR=1.32, 95% CI=1.02-1.72, p-value=0.04). AUC was 0.70, which indicates fair discrimination, although these results should be interpreted with caution due to small sample size (**Figure 4F**).

In comparison, we performed ROC analysis using SORCS3 levels for MDD after adjustment for age and sex in the MPIP MDD sample. This analysis showed 9% increased risk of MDD by 1 ng/ml increase in SORCS3 levels (OR=1.09, 95% CI=1.01-1.18, p-value=0.03). However, AUC was 0.59 indicating no discrimination between patients with and without disease (**Figure 4E**).

## Discussion

To identify genetic loci associated with panic disorder, we carried out a GWAS in the Finnish register-based FinnGen study. We identified significant association with SNPs in the intronic regions of *SORCS3*. We also found higher serum SORCS3 protein levels in panic disorder patients compared to controls. *SORCS3* locus has been linked to other psychiatric endpoints previously, but this is the first report of *SORCS3* associating to panic disorder.

Although there are no previous associations to our lead variants, SNPs that are in LD with them have previously been associated with depression or depressive symptoms ^28, 40–43^, neuroticism ^42, 44–46^, well-being ^42, 44^, risk tolerance ^47^, schizophrenia ^48^, age of smoking initiation ^49^, and gastroesophageal reflux disease ^50^ (**Supplementary Table 1**). Also other genetic regions within the *SORCS3* gene have been associated with a composite phenotype of anxiety disorders ^23^ (the current FinnGen panic disorder sample is included in this PGC-ANX sample), depression ^51^, ADHD ^52^, cognitive abilities ^39, 53, 54^, composite phenotypes consisting of multiple psychiatric and neuropsychiatric disorders ^55^, and Tourette syndrome and comorbid neurodevelopmental disorders in a cross-disorder analysis ^56^. These findings may indicate shared genetic liability, as panic disorder overall correlates genetically with other anxiety disorders ^57^, depression ^15, 58^, and neuroticism ^15^. Of these phenotypes, especially depression is often comorbid with panic disorder, and 43% of panic disorder cases in FinnGen also had a depression diagnosis (**Supplementary Figure 1**). When we excluded all individuals with psychiatric comorbidities, rs902306 and rs1021362 minor alleles still increased risk for panic disorder with similar magnitude as in the analysis of all panic disorder cases. These follow-up analyses suggest that the *SORCS3* lead SNPs are associated with panic disorder in the FinnGen.

We estimated the SNP-based heritability of panic disorder to be 0.14, which is similar in magnitude compared with estimates from other GWASs of mood and anxiety phenotypes ^7, 10, 28^, although somewhat lower than 0.28 previously found for panic disorder specifically ^15^. In general, we found the genetic correlations with other psychiatric phenotypes to be somewhat higher than previously reported ^15^, possibly owing to the decreased sampling error due to the larger sample size. However, in the largest GWAS of anxiety phenotypes to date, the genetic correlations are similar to ours ^23^.

Unexpectedly, our *SORCS3* finding with panic disorder did not replicate in the UK Biobank or Estonian Biobank samples. This may be due to several reasons, including differences in ascertainment and catchment of subjects with panic disorder between biobanks. Also, diagnostic practices differ between the three countries. Although we used very similar inclusion/exclusion criteria for case and control definitions between cohorts, the case fractions were very different (2.2% in the FinnGen, 0.4% in the UK Biobank, and 7.0% in the Estonian Biobank), with the FinnGen estimate being closest to the general lifetime prevalence of panic disorder. It is unlikely that the *SORCS3* association with panic disorder is specific for the Finnish population because the risk allele frequency of rs902306 is similar in the FinnGen subjects (21%) and other European populations (20-27%). The upcoming meta-analysis of panic disorder with considerably larger sample size and cases from multiple populations will help understand whether the lead SNPs identified here and other SNPs within the *SORCS3* locus associate with panic disorder in general.

*SORCS3* belongs to the VPS10P domain gene family, together with *SORCS1*, *SORCS2, SORT1,* and *SORL1*. They are transmembrane proteins that participate in protein transport and signal transduction by binding and internalizing ligands, including trophic factors and neuropeptides ^59^. For example, *SORCS1* has been associated with Alzheimer’s disease ^60^, and in line with the known involvement of amyloid precursor protein (APP) in the pathogenesis of Alzheimer’s disease, both SORCS1 and SORL1 affect processing and transport of APP ^61, 62^. Less is known about the function of SORCS3. Its expression is mostly restricted to the brain ^63, 64^. Simultaneous knockout of *Sorcs1* and *Sorcs3* in mice disturbs hypothalamic control of metabolism through regulation of orexigenic peptide production ^64^. In hippocampus, SORCS3 is located in the same protein complex with PSD-95 and PICK1, where it is suggested to influence postsynaptic AMPA receptor mobility ^65, 66^. Consistent with the putative function of SORCS3 in hippocampal plasticity, *Sorcs3* knockout mice have increased fear memory extinction in an inhibitory avoidance task with normal fear acquisition. These mice also have poor spatial learning and memory in the Barnes maze ^65^. Conclusively, these results suggest that SORCS3 is a regulator of synaptic plasticity and behavior.

Like other VPS10P domain family members, SORCS3 protein’s extracellular domain can be shed by an enzymatic cleavage ^67^. The functional significance of this shedding remains unclear. However, SORCS3 may function as a carrier protein for its ligands, such as the platelet-derived growth factor BB ^67^, nerve growth factor ^68^, or tropomyosin-related kinase B ^64^. We measured the concentration of the shed SORCS3 protein in the serum of MPIP panic disorder and MDD patient and control samples from Germany. We found 41% higher SORCS3 levels in panic disorder patients compared to controls. In the WU replication sample, SORCS3 serum levels were 30% higher in panic disorder patients compared to controls. Furthermore, ROC curve analysis suggested serum SORCS3 levels to have the ability to diagnose patients with and without panic disorder, but not MDD. However, SORCS3 plasma levels did not differ between panic disorder or MDD cases and controls in a Finnish biobank sample. The SORCS3 levels were significantly lower in plasma than serum, and 35% of the samples were below the detection limit of the ELISA kit. This may be one reason for the lack of replication. SORCS3 serum levels have not been previously associated with clinical phenotypes, and its source remains unclear. SORCS3 expression is highest in the brain, but lower expression levels are also detectable in the adrenal glands and peripheral nerves according to the Genotype-Tissue Expression Project. Of the other sortilin family members, serum SORT1 levels were shown to be 11.5% higher in patients of MDD compared to controls ^69^. Peripheral biomarkers for the diagnosis, prognosis, and treatment response of panic disorder are greatly needed. Our data suggest SORCS3 may be a putative serum biomarker, and investigation of its function in panic disorder may reveal new pathogenetic mechanisms.

The main strength of the study is the biobank-based sample from a genetically homogeneous population that allowed sample size sufficient to identify a novel risk locus for panic disorder. Also, we were able to use lifetime diagnoses of anxiety disorders from nationwide health registers. These health register-based diagnoses are also the main limitation of the study. Finns suffering from anxiety disorders contact health services more often than the rest of the population ^70^ and are thus well represented within health registers. However, only 33.5-70.9 % of individuals meeting diagnostic criteria for an anxiety disorder contact health care services specifically for mental health reasons ^70, 71^, and therefore may not necessarily get an anxiety disorder diagnosis. Although we excluded individuals with any psychiatric diagnoses as controls, our controls may include individuals with genetic liability to panic disorder. Thus, our results may not be generalizable to the wider population or all individuals with panic disorder. Larger studies to identify additional and distinct loci for panic disorder are needed in the future to confirm our findings.

In conclusion, genetic variants within introns 1 and 2 of the *SORCS3* gene associated significantly with panic disorder in the FinnGen. *SORCS3* has previously been associated with other psychiatric disorders, such as a composite anxiety disorder phenotype and MDD. We also found SORCS3 serum levels to be significantly higher in panic disorder patients compared to controls, suggesting that SORCS3 may serve as a novel biomarker for panic disorder. The functions of SORCS3 in the brain and blood are largely unknown. Revealing the neurobiological mechanisms by which SORCS3 regulates anxiety may help understand the etiology of panic disorder.

## Data Availability

All data produced in the present study are available upon reasonable request to the authors

## Acknowledgements

We thank the FinnGen analysis team, especially Timo Sipilä, for help with data analysis. This work was funded by the Sigrid Jusélius Foundation (IH), Max Planck Society (EB, CWT, BN, AEL), German Research Foundation (AP, RP, JBL) and the EU ERDF grant no. 2014-2020.4.01.15-0012 (MV, AM, Estonian Center of Excellence in Genomics and Translational Medicine). The UKBB replication study has been conducted using the UK Biobank Resource under application number 31063. The FinnGen project is funded by two grants from Business Finland (HUS 4685/31/2016 and UH 4386/31/2016) and the following industry partners: AbbVie Inc., AstraZeneca UK Ltd, Biogen MA Inc., Bristol Myers Squibb (and Celgene Corporation & Celgene International II Sàrl), Genentech Inc., Merck Sharp & Dohme LCC, Pfizer Inc., GlaxoSmithKline Intellectual Property Development Ltd., Sanofi US Services Inc., Maze Therapeutics Inc., Janssen Biotech Inc, Novartis AG, and Boehringer Ingelheim International GmbH. Following biobanks are acknowledged for delivering biobank samples to FinnGen: Auria Biobank (www.auria.fi/biopankki), THL Biobank (www.thl.fi/biobank), Helsinki Biobank (www.helsinginbiopankki.fi), Biobank Borealis of Northern Finland (https://www.ppshp.fi/Tutkimus-ja-opetus/Biopankki/Pages/Biobank-Borealis-briefly-in-English.aspx), Finnish Clinical Biobank Tampere (www.tays.fi/en-US/Research_and_development/Finnish_Clinical_Biobank_Tampere), Biobank of Eastern Finland (www.ita-suomenbiopankki.fi/en), Central Finland Biobank (www.ksshp.fi/fi-FI/Potilaalle/Biopankki), Finnish Red Cross Blood Service Biobank (www.veripalvelu.fi/verenluovutus/biopankkitoiminta) and Terveystalo Biobank (www.terveystalo.com/fi/Yritystietoa/Terveystalo-Biopankki/Biopankki/). All Finnish Biobanks are members of BBMRI.fi infrastructure (www.bbmri.fi). Finnish Biobank Cooperative -FINBB (https://finbb.fi/) is the coordinator of BBMRI-ERIC operations in Finland. The Finnish biobank data can be accessed through the Fingenious^®^ services (https://site.fingenious.fi/en/) managed by FINBB. We also acknowledge the Helsinki Biobank (www.helsinginbiopankki.fi) for plasma samples used for measuring SORCS3 levels.

