## Supplemental information for "Genome-wide Association Study Identifies *SORCS3* as a Novel Susceptibility Locus for Panic Disorder in the FinnGen Study"

### Supplemental Methods

**The FinnGen Study Sample**

The FinnGen (https://www.finngen.fi/en) study combines genotype data with longitudinal health register data of Finland. FinnGen study participants were recruited from epidemiological and disease-based cohorts, as well as from hospital biobanks. We used the FinnGen Data Freeze 6, with a total of 260,405 individuals (147,061 females and 113,344 males) and register data deriving from the Causes of Death (from 1969), Inpatient Care (from 1969 to 1993) and Specialized Outpatient Care (from 1994), Surgeries and Outpatient Care (from 1998), Primary Health Care (2011-2018), and Drug Reimbursement (from 1964) Registers. All FinnGen data is pseudonymized.

Patients and control subjects in FinnGen provided informed consent for biobank research, based on the Finnish Biobank Act. Alternatively, separate research cohorts, collected prior the Finnish Biobank Act came into effect (in September 2013) and start of FinnGen (August 2017), were collected based on study-specific consents and later transferred to the Finnish biobanks after approval by Fimea (Finnish Medicines Agency), the National Supervisory Authority for Welfare and Health. Recruitment protocols followed the biobank protocols approved by Fimea. The Coordinating Ethics Committee of the Hospital District of Helsinki and Uusimaa (HUS) statement number for the FinnGen study is Nr HUS/990/2017. The FinnGen study is approved by Finnish Institute for Health and Welfare (permit numbers: THL/2031/6.02.00/2017, THL/1101/5.05.00/2017, THL/341/6.02.00/2018, THL/2222/6.02.00/2018, THL/283/6.02.00/2019, THL/1721/5.05.00/2019 and THL/1524/5.05.00/2020), Digital and population data service agency (permit numbers: VRK43431/2017-3, VRK/6909/2018-3, VRK/4415/2019-3), the Social Insurance Institution (permit numbers: KELA 58/522/2017, KELA 131/522/2018, KELA 70/522/2019, KELA 98/522/2019, KELA 134/522/2019, KELA 138/522/2019, KELA 2/522/2020, KELA 16/522/2020), Findata permit numbers THL/2364/14.02/2020, THL/4055/14.06.00/2020,,THL/3433/14.06.00/2020, THL/4432/14.06/2020, THL/5189/14.06/2020, THL/5894/14.06.00/2020, THL/6619/14.06.00/2020, THL/209/14.06.00/2021, THL/688/14.06.00/2021, THL/1284/14.06.00/2021, THL/1965/14.06.00/2021, THL/5546/14.02.00/2020, THL/2658/14.06.00/2021, THL/4235/14.06.00/202, Statistics Finland (permit numbers: TK-53-1041-17 and TK/143/07.03.00/2020 (earlier TK-53-90-20) TK/1735/07.03.00/2021, TK/3112/07.03.00/2021) and Finnish Registry for Kidney Diseases permission/extract from the meeting minutes on 4^th^ July 2019. The Biobank Access Decisions for FinnGen samples and data include: THL Biobank BB2017_55, BB2017_111, BB2018_19, BB_2018_34, BB_2018_67, BB2018_71, BB2019_7, BB2019_8, BB2019_26, BB2020_1, Finnish Red Cross Blood Service Biobank 7.12.2017, Helsinki Biobank HUS/359/2017, HUS/248/2020, Auria Biobank AB17-5154 and amendment #1 (August 17 2020), AB20-5926 and amendment #1 (April 23 2020) and it´s modification (Sep 22 2021), Biobank Borealis of Northern Finland_2017_1013, Biobank of Eastern Finland 1186/2018 and amendment 22 § /2020, Finnish Clinical Biobank Tampere MH0004 and amendments (21.02.2020 & 06.10.2020), Central Finland Biobank 1-2017, and Terveystalo Biobank STB 2018001 and amendment 25^th^ Aug 2020.

Cases were defined according to ICD-10 F41.0 as having panic disorder. Additionally, we included subjects diagnosed using ICD-9 3000B (panic disorder) as cases – DSM-III-R diagnostic criteria were used in Finland for ICD-9. The Finnish ICD system allowed us to classify the cases to mild, moderate, or severe panic disorder (F41.08, F41.00, F41.01), or unspecified panic disorder (F41.09) (see **Table 1**). We excluded cases with psychotic disorder, autism, or intellectual disability (ICD-10 F20-F29, F70-F79, F84.0, F84.1, F84.5; ICD-9 295, 297, 298, 2990, 317-319; ICD-8 295, 297, 298, 29999, 311-315). Exclusion criteria for controls included history of any psychiatric endpoint (ICD-10 F00-F99, ICD-9 290-319, or ICD-8 290-315). Additionally, we adjusted the age range of controls to match to that of cases. Based on this approach, we included 3,549 cases and 159,869 control subjects for our study, yielding a case fraction of 2.2% representing panic disorder in the general Finnish population^1^. We used a similar approach for the comparative analyses with other psychiatric endpoints while defining cases and controls for major depression disorder (MDD) and general anxiety disorder (GAD) including phobias. More specifically, MDD cases were defined as having been diagnosed according to ICD-10 (F32, F33); ICD-9 (2961); or ICD-8 (2980, 3004, 79020) codes. GAD and phobia cases were individuals positive for ICD-10 F40.00, F40.01, F40.1, F40.2, F40.8, F40.09, F41.1, F41.2, F41.3, F41.8, F41.9; ICD-9 3000C, 3002C, 3002D, 3002X; or ICD-8 300.00 or 300.20.

**The UK Biobank Study Sample**

Participants between 40-69 years of age who were registered with a general practitioner of the UK National Health Service (NHS)^2^ were recruited. From 2006-2010, a total of 503,325 individuals were included. All study participants provided informed consent and the study was approved by the North West Multi-centre Research Ethics Committee. Cases were defined according to ICD-10 codes as having panic disorder (F41.0). Individuals with psychotic disorder, autism, or intellectual disability from cases (ICD-10 F20-F29, F70-F79, F84.0, F84.1, F84.5; ICD-9 295, 297, 298, 2990, 317-319) Control subjects were free from any psychiatric diagnoses (ICD-10 F00-F99, ICD-9 290-319). In contrast to filtering applied in the FinnGen sample, no age restriction was imposed on the control group as the age distributions between the cases and controls are very similar. After applying the above inclusion/exclusion criteria, 1,531 cases and 358,035 controls with European ancestry were available for replication analysis with *SORCS3* lead variants. It is notable that based on this data, case fraction in the UK Biobank sample for replication analyses was 0.4%, which is ~4 times lower than estimated 1.7% prevalence of panic disorder in the UK^3^. GWAS was carried out based on a logistic mixed model with first 10 genetic PCs, age, age^2^, sex, age $\times$ sex, age^2^ $\times$ sex as covariates using SAIGE v. 0.44.1^4^, which accounts for sample relatedness and the unbalanced case-control ratio for panic disorder. SNPs with a MAC < 20 and/or imputation score < 0.3 were excluded from the results.

**The Estonian Biobank Study Sample**

The panic disorder diagnoses were derived from digital health registries and have been mainly diagnosed by psychiatrists. In Estonia, digital health registries have been used since 1999. According to the Estonian Health Statistics and Health Research Database, from 1999 to 2020, 3.17% of the Estonian population has been diagnosed with panic disorder (2.1% of men, 4.15% of women)^5^. Estonian Biobank (EstBB) uses information from the same digital registries and during 1999-2020, 3.5% of the EstBB sample has been diagnosed with panic disorder (2.11% of men, 4.27% of women)^5^, thus, the prevalence of panic disorder is similar in the general population and in the EstBB sample. Cases were defined according to ICD-10 codes as having panic disorder (F41.0). Cases with psychotic disorder, autism, or intellectual disability (ICD-10 F20-F29, F70-F79, F84.0, F84.1, F84.5) were excluded from both cases and controls. In addition, the exclusion criteria for controls included history of any psychiatric endpoint (ICD-10 F00-F99) and the age range of controls was adjusted to match that of cases. The EstBB sample for replication analyses included 7,057 cases (1,455 males and 5,602 females) and 94,024 controls (37,119 males and 56,905 females) yielding case fraction of 7.0%, which is ~2 times the prevalence of panic disorder in EstBB overall or in the Estonian population. GWAS for panic disorder was carried out using SAIGE v. 0.43.1 ^4^ after adjustment for PC1-10, age, and sex, and excluding SNPs with a MAF < 0.1% and/or imputation score < 0.4 were removed from the results.

**The Max Planck Institute of Psychiatry Study Sample**

Panic disorder patients were recruited in the Anxiety Disorders Outpatient Unit at the Max Planck Institute of Psychiatry (MPIP) in Munich. Panic disorder with and without agoraphobia was the primary diagnosis, mild and moderate secondary depressive symptoms were allowed. The diagnosis was ascertained by trained psychiatrists according to the Diagnostic and Statistical Manual of Mental Disorders (DSM)-IV criteria. All patients underwent the Structured Clinical Interviews for DSM-IV (SCID I and II). Panic disorder due to a medical or neurological condition or the presence of a comorbid Axis II disorder was an exclusion criterion. All patients underwent a thorough medical examination including EEG, ECG, and detailed hormone laboratory assessment [for more detailed description see ^6^]. Control subjects were recruited from a Munich-based community sample and screened for the absence of axis I psychiatric disorders using the Munich version of the Composite International Diagnostic Interview (M-CIDI)^7^, which specifically screen for alcohol dependence, drug abuse or dependence, possible psychotic disorder, mood disorder, anxiety disorder including OCD and PTSD, somatoform disorder, dissociative disorder NOS, and eating disorder. Controls were age- and sex-matched with patients. All subjects were Caucasian and provided written informed consent. The study was approved by the ethics committee of Ludwig-Maximilians-University in Munich, Project number 318/00 and was designed as well as performed in accordance with the Declaration of Helsinki. Written informed consent was obtained from all subjects.

An independent depression sample (n=102) was recruited at the MPIP within the Munich Antidepressant Respose Study (MARS). Recruitment strategies and detailed characterization of participants for the whole sample have been described elsewhere ^8, 9^. In short, the diagnosis was ascertained by trained psychiatrists according to the DSM-IV criteria. The exclusion criteria were the presence of alcohol or substance abuse or dependence, comorbid somatization disorder, and depressive disorders owing to general medical or neurologic conditions. All patients were medicated. Healthy controls were recruited and characterized in the same way as described in the section for panic disorder. The study was approved by the local ethics committee and all individuals gave written informed consent.

**Würzburg Study Sample (WU)**

Patient samples: The current study is part of an overarching clinical research project in which an exposure-based cognitive behavioral therapy for anxiety disorders was complemented by laboratory assessments before and after treatment. For this project, patients with anxiety disorders aged between 18 and 65 years were recruited via the outpatient clinic of the University of Würzburg, local advertisement, and referral. Inclusion criteria required a primary diagnosis of social anxiety disorder, agoraphobia, or panic disorder. Exclusion criteria comprised acute suicidality, psychosis, bipolar disorder, borderline personality disorder, substance dependence, intellectual disability, any severe acute medical condition, pregnancy, medical advice to avoid stressful situations, and any contraindication for exposure therapy. Other comorbidities and intake of psychiatric medication were not considered exclusion criteria. Diagnostic interview for mental disorders was conducted using the Mini-DIPS, a structured clinical interview assessing anxiety, affective, obsessive-compulsive, trauma-related, eating and several other mental disorders according to DSM-5 ^10^. However, if applicable, medication had to be taken for at least 3 months with a stable dose during the last 4 weeks (see also detailed description in ^11^). Bioprobes were collected on the day of the laboratory assessment before and after the CBT intervention (12 sessions exposure-based CBT) as well as 6 months later. All participants provided written informed consent to the procedures approved by the local ethics committee (GZEK2018‐20).  In total, serum samples from 24 panic disorder patients with or without agoraphobia (18 females and 6 males) were analyzed in this study.

Control samples: Participants were recruited and assessed at the Center of Mental Health, Department of Psychiatry, Psychosomatics and Psychotherapy, University Hospital of Würzburg, Germany within the DFG-funded study investigating fear generalization (Project number 499262975). All participants provided written informed consent prior to participation. Inclusion criteria for the current analysis was absence of any mental illness assessed by the Mini-DIPS ^10^, conducted by a clinical psychologist. Serious neurological and somatic illnesses as well as pregnancy were exclusion criteria. Blood samples including serum were collected following the experimental paradigm to analyze blood cell composition and routine indices. Samples were processed immediately after collection and stored at -80°C for later analysis. The project has been approved by the local ethical committee (256/21).

**Helsinki Biobank Study Sample**

Plasma samples from 300 panic disorder patients, 300 patients with major depressive disorder, and 300 controls were obtained from the Helsinki Biobank. Inclusion criterion for panic disorder was ICD-10 F41.0 diagnosis and exclusion criteria psychotic disorders (F20-F29), autism (F84.0, F84.1, F84.5), or intellectual disability (F70-F79). Inclusion criterion for major depressive disorder was ICD-10 F33 diagnosis and exclusion criteria psychotic disorders (F20-F29), autism (F84.0, F84.1, F84.5), or intellectual disability (F70-F79) and panic disorder. We required that the plasma sample was taken after the first diagnosis (the average time being 6.8 years). Exclusion criterion for controls included history of any psychiatric endpoint. Additionally, we matched sex and age of the controls to both case groups. The study was approved by the Helsinki University Hospital Ethics Committee (22/2023).

### Supplemental Table 1. PheWAS Results with SORCS3 Lead and Proxy SNPs.

| **Lead SNP** | **Chr** | **Pos (hg38)** | **Proxy SNP** | **Proxy pos (hg38)** | **Distance (bp)** | **r2** | **Trait** | **p-value** | **PMID** |
| --- | --- | --- | --- | --- | --- | --- | --- | --- | --- |
| **rs902306** | 10 | 104,772,457 | rs1490176 | 104,800,467 | -28,010 | 0.87 | Neuroticism | 1x10^-12^ | 30643256 |
|  |  |  |  |  |  |  | Positive affect | 2x10^-12^ | 30643256 |
|  |  |  |  |  |  |  | Life satisfaction | 1x10^-11^ | 30643256 |
|  |  |  |  |  |  |  | Depressed affect | 2x10^-08^ | 29942085 |
|  |  |  | rs12416372 | 104,698,869 | 73,588 | 0.85 | General factor of neuroticism | 7x10^-09^ | 30867560 |
|  |  |  | rs2491392 | 104,798,186 | -25,729 | 0.80 | General risk tolerance (MTAG) | 2x10^-08^ | 30643258 |
| **rs1021362** | 10 | 104,851,510 | rs1021363 | 104,851,081 | 429 | 1 | Depression | 4x10^-23^ | 30718901 |
|  |  |  |  |  |  |  | Major depressive disorder (MTAG) | 2x10^-14^ | 33479212 |
|  |  |  |  |  |  |  | Depressive symptoms | 1x10^-13^ | 30643256 |
|  |  |  |  |  |  |  | Neuropsychiatric disorders | 4x10^-12^ | 33479212 |
|  |  |  |  |  |  |  | Gastroesophageal reflux disease | 5x10^-10^ | 34187846 |
|  |  |  |  |  |  |  | Autism spectrum disorder (MTAG) | 1x10^-08^ | 33479212 |
|  |  |  |  |  |  |  | Depression (broad) | 1x10^-08^ | 29662059 |
|  |  |  |  |  |  |  | Major depressive disorder | 5x10^-25^ | 37464041 |
|  |  |  | rs2496024 | 104,876,838 | -25,328 | 0.95 | Age at first sexual intercourse | 3x10^-21^ | 34211149 |
|  |  |  |  |  |  |  | Cognitive performance (MTAG) | 5x10^-14^ | 30038396 |
|  |  |  |  |  |  |  | Depressive symptoms (MTAG) | 5x10^-13^ | 32606422 |
|  |  |  |  |  |  |  | Lifetime smoking index | 1x10^-11^ | 31689377 |
|  |  |  |  |  |  |  | Smoking status | 7x10^-10^ | 30595370 |
|  |  |  |  |  |  |  | Childhood maltreatment | 7x10^-09^ | 33740410 |
|  |  |  |  |  |  |  | Intelligence | 3x10^-08^ | 29942086 |
|  |  |  | rs2496022 | 104,872,720 | -21,210 | 0.95 | Depressive symptoms (MTAG) | 4x10^-14^ | 29292387 |
|  |  |  |  |  |  |  | Depressive symptoms | 7x10^-11^ | 29292387 |
|  |  |  | rs1961639 | 104,876,239 | -24,729 | 0.95 | Subjective well-being (MTAG) | 2x10^-12^ | 29292387 |
|  |  |  |  |  |  |  | Depressive symptoms | 4x10^-09^ | 32606422 |
|  |  |  |  |  |  |  | Depression | 4x10^-09^ | 29942085 |
|  |  |  | rs2451500 | 104,873,119 | -21,609 | 0.95 | Neuroticism (MTAG) | 6x10^-11^ | 29292387 |
|  |  |  | rs2491383 | 104,856,882 | -5,372 | 0.82 | Age of smoking initiation (MTAG) | 1x10^-10^ | 30643251 |
|  |  |  | rs10786831 | 104,854,813 | -3,303 | 0.82 | Major depressive disorder | 8x10^-09^ | 27479909 |
|  |  |  | rs7900775 | 104,840,585 | 10,925 | 0.93 | Schizophrenia | 4x10^-09^ | 33169155 |

### Supplemental Table 2. Comparative SORCS3 SNP Associations with Other Psychiatric Disorders in the FinnGen.

|  | | | **rs902306** | | **rs1021362** | |
| --- | --- | --- | --- | --- | --- | --- |
| **Phenotype** | **Cases** | **Controls** | **OR (95% CI)** | **p-value** | **OR (95% CI)** | **p-value** |
| ^a^Panic disorder only | 628 | 159,869 | 1.20 (1.04-1.37) | 0.01 | 1.24 (1.10-1.41) | 6.3x10^-04^ |
| ^b^GAD/phobia | 3,738 | 161,438 | 1.07 (1.01-1.13) | 0.03 | 1.03 (0.98-1.09) | 0.25 |
| ^b^MDD | 25,906 | 161,371 | 1.05 (1.03-1.08) | 6.4x10^-05^ | 1.05 (1.02-1.07) | 6.6x10^-05^ |

GAD, generalized anxiety disorder; MDD, major depressive disorder, OR, odds ratio; CI, confidence interval.

^a^Panic disorder only cases are free from any psychiatric comorbidities, such as MDD, GAD/phobia, or other anxiety or stress disorder.

^b^Panic disorder is excluded from GAD/phobia or MDD case definition.

### Supplemental Table 3. Replication of Lead SORCS3 Variants with Panic Disorder in the UK Biobank and Estonian Biobank.

|  |  |  |  | rs902306 |  | rs1021362 | | |
| --- | --- | --- | --- | --- | --- | --- | --- | --- |
| Study | Cases | Controls | EAF | OR (95% CI) | p-value | EAF | OR (95% CI) | p-value |
| UK Biobank | 1,531 | 358,035 | 0.26 | 1.00 (0.92-1.09) | 0.99 | 0.36 | 1.01 (0.94-1.09) | 0.75 |
| Estonian Biobank | 7,057 | 94,024 | 0.26 | 1.02 (0.98-1.07) | 0.25 | 0.35 | 1.00 (0.96-1.03) | 0.82 |

EAF, effect allele frequency; OR, odds ratio; CI, confidence interval.

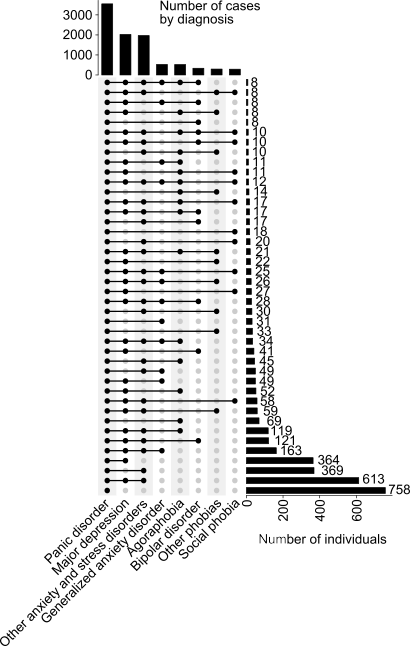

**Supplemental Figure 1. Panic disorder comorbidities within FinnGen**. The bars on the right show the numbers of individuals with the specific comorbidity indicated by the dots on the left side (e.g. the bottom row refers to individuals with a panic disorder diagnosis, but no other diagnoses from the other categories). The bars on the top show overall number of individuals within each diagnostic category. The ICD-codes for categories not described elsewhere in the Materials and Methods section are as follow:

GAD: ICD-10: F41.1; ICD-9: 3000C

Agoraphobia: ICD-10: F40.0; ICD-9: 3002C

Bipolar disorder: ICD-10: F31; ICD-9: 2961, 2963, 2964, 2967

Other phobias: ICD-10: F40.2, F40.8, F40.9; ICD-9: 3002X; ICD-8: 300.20

Social phobia: ICD-10: F40.1; ICD-9: 3002D

Other anx & stress (captures any anxiety or stress related disorder not otherwise included): ICD-10: F41.2, F41.3, F41.8, F41.9, F42, F43, F44, F45, F48; ICD-9: 3000A, 3001, 3003, 3004, 3006, 3007, 3008, 3009; ICD-8: 300.00, 300.1, 300.3, 300.4, 300.5, 300.6, 300.7, 300.8, 300.9)

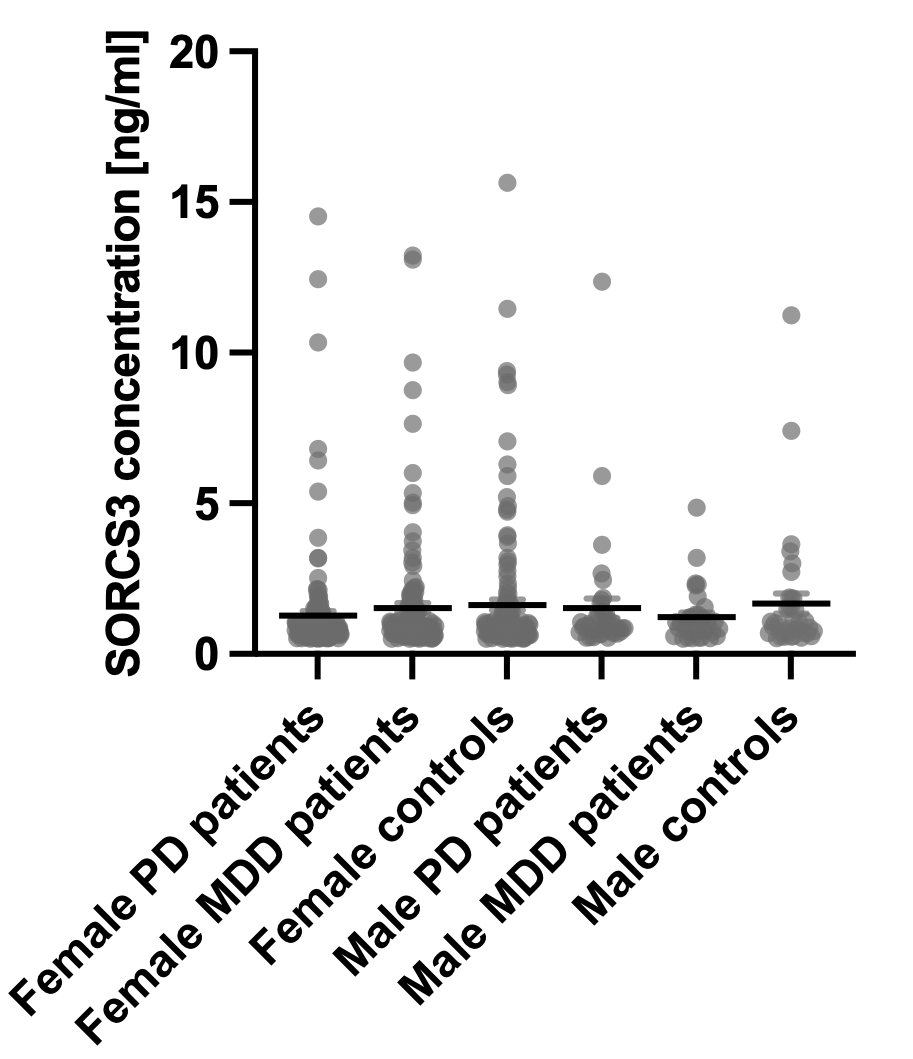

**Supplemental Figure 2. Plasma SORCS3 levels in patients with panic disorder (PD), major depressive disorder (MDD) and controls.** SORCS3 protein concentration was measured by ELISA from samples obtained from the Helsinki Biobank. Only samples where the concentration was within the detection limit of the kit (0.5-16 ng/ml) are included (n=242 female PD patients, n=246 female MDD patients, n=242 female controls, n=58 male PD patients, n=54 male MDD patients, n=58 male controls). Mean +/- SEM is shown in addition to the individual data points.
